# Individual and spatial heterogeneity of praziquantel efficacy against *Schistosoma mansoni* within the context of repeated mass drug administration

**DOI:** 10.64898/2026.01.04.25342566

**Authors:** Melissa A. Iacovidou, Fabian Reitzug, Sophie Winter, Annet Enzaru, Emily Asiimwe, Juliet Nambatya, Aisha Nakato, Moses Semakula, Betty Nabatte, Narcis B. Kabatereine, Goylette F. Chami

## Abstract

**Background:** Schistosomiasis control relies on the continued effectiveness of praziquantel (PZQ), yet individual and spatial heterogeneity in PZQ efficacy in the context of repeated mass drug administration (MDA) remains poorly understood. This study aimed to identify individual and spatial determinants of PZQ efficacy against *Schistosoma mansoni* in rural Uganda.

**Methods:** We studied 3870 participants aged 5–90 years from 52 villages in Pakwach, Buliisa, and Mayuge districts in Uganda. Participants were recruited to the SchistoTrack cohort in January–February of 2022 and 2023. Participants received PZQ and were followed up four to five weeks later to assess cure using Kato—Katz (KK) microscopy and point-of-care circulating cathodic antigen tests. Logistic regression models were used to identify predictors of cure, defined as egg reduction rates (ERR) of 100% or ERR ***≥***90%. We explored a comprehensive set of 18 sociodemographic, biomedical, water, sanitation, and hygiene, and spatial factors. Subgroup analyses were conducted for adults (aged 18–90) versus children (aged 5–17). Spatial autocorrelation in the outcome and model residuals was assessed using join count statistics and Moran’s *I*.

**Results:** Of 3870 clinical participants, 3704 (95.7%) received PZQ, and 3395 of these had complete clinical data. Among the 3395 treated participants, 1406 (41.4%) were infected with *S. mansoni* at baseline. The overall cure rate (ERR of 100%) was 76.3%, ranging from 68.9% to 85.4% across districts. Higher odds of cure were associated with older age (OR 1.34) and lower baseline infection intensity (moderate/heavy vs light OR 0.39–0.61). Greater height was associated with lower odds of cure (OR 0.99), which was driven by adults. Compared with Mayuge district, participants in Western districts had lower odds of cure (OR 0.48–0.53). In children, greater distance to water sites with snail presence was linked to higher odds of cure (OR 1.05 per 100 m). Results were similar when defining cure as ERR ***≥***90% and when using POC-CCA outcomes. Spatial clustering of treatment outcomes was observed by district, but no residual spatial autocorrelation remained after accounting for district effects.

**Conclusions:** Incorporating spatial analyses, refining dose assessment, and strengthening post-MDA monitoring may help maintain PZQ effectiveness and support sustainable schistosomiasis control efforts.

## Background

Regular monitoring of mass drug administration (MDA) programmes is essential to sustain progress towards schistosomiasis control and elimination targets set by the World Health Organization (WHO). These targets include the elimination of schistosomiasis as a public health problem in all 78 endemic countries and interruption of human transmission in 25 selected endemic countries by 2030 [1]. The main control strategy is through repeated MDA with praziquantel (PZQ), and relies on its continued effectiveness across endemic regions. There is high heterogeneity in drug responses for individuals. Meta-analyses of PZQ efficacy have shown high but variable average cure rates ranging from 73.6% to 94.7% and egg reduction rates (ERRs) ranging from 68.2% to 99.9% across geographies, with particular low efficacy for *Schistosoma mansoni* species [2–6]. Although there is no confirmed evidence of stable PZQ resistance in human populations [6], recent genomic studies have identified loci potentially associated with reduced sensitivity to PZQ [7, 8]. There remains a need to understand sociodemographic, history of treatment, and spatial factors that drive observed variation in treatment outcomes.

There is limited information on the influence of sociodemographic factors beyond age and gender on drug efficacy. Most field studies evaluating PZQ efficacy have focused on children, most commonly school-aged but also preschool-aged, in limited geographic areas [3, 9, 10]. Sustained schistosome exposure and an observed high burden of morbidity in adults, including regional variation in both exposure and morbidity [11, 12], warrant a more detailed assessment across larger age ranges, especially in regards to treatment and monitoring [13]. There is no evidence on gender differences in treatment response, with studies reporting no association between gender and cure rates or ERR [9, 14–20]. Important gaps also remain in understanding if and how other socioeconomic determinants, including education level and occupation, as well as water, sanitation, and hygiene (WASH) conditions, influence drug efficacy; while these factors are well-recognised contributors to schistosomiasis risk [21, 22], they are rarely assessed in PZQ efficacy studies.

History of treatment may also contribute to observed variation in PZQ response. Modelling studies have incorporated treatment history to assess the cumulative impact of repeated exposure to PZQ, typically in the form of rounds of MDA reported in the area, without considering adherence or the number of treatments received in a defined period of time [16, 17]. Crellen et al. reported that repeated rounds of MDA were associated with reduced PZQ efficacy [17], and called for close monitoring of PZQ efficacy.

Spatial heterogeneity in *Schistosoma* populations may influence drug efficacy and contribute to inconsistent treatment outcomes between communities. Country-level differences in drug response are evident by wide ranges of cure rates [4, 23], yet regional and community-level differences have not been adequately explored, other than at the school level for children-only studies [17]. It has been suggested that treatment requirements should account for spatial variation in schistosomiasis risk and avoid country-level aggregation in countries with a heterogeneous distribution of infection [24]. Despite evidence of heterogeneity, transmission models often assume uniform drug response across settings and individuals, potentially oversimplifying the dynamics of control [25–28].

We conducted a comprehensive epidemiological assessment of drug efficacy in rural Uganda. Sociodemographic, biomedical, water, sanitation, and hygiene (WASH), human water contact, and spatial factors were studied in individuals aged 5–90 years from 52 villages in three districts within the community-based cohort, SchistoTrack. We investigated the determinants of being cured after treatment as well as achieving an ERR of 90%. This study aimed to identify whether individual and spatial heterogeneity are present in PZQ efficacy against *Schistosoma mansoni*.

## Methods

### Study design and participants

This study was conducted within the SchistoTrack cohort in the districts of Mayuge, Buliisa, and Pakwach in rural Uganda [29]. The study area has a history of MDA dating back to 2003, with individuals five years or older being treated through both community-based and school-based administration. The last round of MDA before 2022 was in 2020, with Buliisa and Pakwach having participated in 13 rounds while Mayuge received 15 [11]. From the initial 38 villages in 2022 and an additional 14 villages enrolled in 2023 (from Buliisa and Pakwach only), a total of 3870 clinical participants were recruited from a total of 1938 households. Participants included adults and children aged five years or older, aiming for one child and one adult per household.

Details on the study design and participant sampling are provided in Puthur et al. [30]. Follow-up surveys were performed on all newly recruited participants (all participants in 2022 and participants from new villages in 2023) approximately four to five weeks after treatment to assess drug efficacy.

### Outcomes

The primary outcome of interest was whether individuals were cured after treatment. All participants were offered treatment with PZQ regardless of their infection status, administered by government nurses based on the WHO dose pole [31]. Treatment was directly observed and followed by the administration of food and drink. Participants were not treated with PZQ if they had a history of seizures, vomiting blood, were pregnant at the time of survey, presented varices in sonography scans, or showed signs of severe illness. Participants who did not attend clinical surveys at both baseline and follow-up were excluded from the analyses.

All clinical participants were tested for *S. mansoni* infections using Kato–Katz (KK) microscopy on two slides from a single stool sample, which were read by independent technicians, as described elsewhere [11]. Eggs per gram (EPG) values were calculated by averaging across slides and multiplying by a factor of 24. The ERR was calculated as follows:

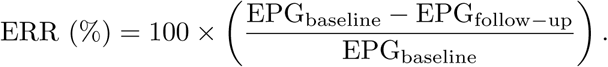

We considered a binary outcome where one denotes 100% ERR, and zero denotes *<* 100%, indicating cured and non-cured individuals, respectively. We also considered a threshold of 90% for the cured outcome, based on the WHO classification criteria whereby *≥* 90% ERR is considered satisfactory [32]. As a secondary outcome, we also considered point-of-care circulating cathodic antigen (POC-CCA) tests, where individuals who tested negative at follow-up were marked as cured (see Supplementary methods). We considered tests classified as trace (barely visible treatment line) to be positive (POC-CCA+) or negative (POC-CCA*−*) in separate models.

### Covariates

The covariates of interest included sociodemographic, biomedical, WASH, water contact, spatial, and other study design factors. These were chosen based on their importance in the literature and their representation of infection exposure (e.g. water contact), potential approximation of acquired immunity (e.g. age), MDA treatment history, or possible influence on drug metabolism (e.g. age, height/weight).

Five sociodemographic and economic variables were measured. Age (in years) was recorded at the time of recruitment. Gender was a binary variable with female coded as one. Educational attainment was coded numerically to indicate the highest level of education completed and ranged from 0 to 14, with the lowest corresponding to no education and the highest representing completion of university education, as defined elsewhere [11, 30]. The main income-earning occupation (if any) for each individual was coded as a categorical variable to represent occupations with high schistosome exposure [12], including fishermen and fishmongers, with all other occupations and no occupation being the reference. The number of years the household has been in the village was included as a proxy indicator, along with age, for acquired immunity and treatment history.

Four biomedical variables were observed. Baseline current schistosome infection intensity was coded as none, light (1–99 EPG), moderate (100–399 EPG), and heavy (*≥* 400 EPG), as per the WHO guidelines [1]. Nurses asked whether the participants had received any PZQ within the last year outside of the study to assess participation in MDA, and this was included as a binary variable with one representing MDA participation. This variable was also used as a proxy indicator of the history of MDA treatment, as shown elsewhere [33]. A second set of nurses conducted anthropometric measurements of height (nearest cm) and weight (nearest 0.1 kg), using tape measures fixed to walls and manual scales (Elite weighing scales and DT01 mechanical personal scales) placed on flat surfaces. PZQ dosage is based on weight, where dosage is recommended at 40 mg*/*kg [31]. Height is used for the WHO dose pole as a direct approximation of weight [34]. We also computed the body mass index (BMI) of participants 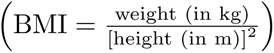 to investigate whether height was an adequate proxy for weight, motivated by concerns raised in a recent paper reviewing the dose pole [35]. BMI categories for adults were defined as per the WHO classification [36]: underweight (*<* 18.5 kg*/*m^2^), normal (18.5 to 24.9 kg*/*m^2^), overweight (*≥* 25.0 kg*/*m^2^), and obese (*≥* 30.0 kg*/*m^2^).

Detailed information on WASH and human water contact was collected to understand the history of exposure and past likelihood of reinfection. Improved sanitation was defined as having access to a flush/pour-flush toilet or a private covered latrine using the United Nations Children’s Fund (UNICEF)/WHO definition of WASH indicators [37]. This variable was included due to its relevance to infection and influence on environmental risk [22]. Self-reported participation in different water contact activities was used as a proxy of schistosome exposure, where activities were grouped in three broader categories, as per Reitzug et al. [12]: 1) domestic, including getting drinking water, washing clothes, bathing, and washing jerry cans or household items; 2) occupational, including collecting papyrus, fishing, fishmongering, and collecting shells; and 3) recreational, including swimming or playing in water. These activities have been shown to have a high association with increased infection risk [38].

The distance to the nearest water site with snail presence from the household of the participant (m) was included to represent the environmental risk of schistosome infection [39]. Furthermore, the distance to the nearest government health centre from the household of the participant (km) was included as a proxy indicator for access to care more generally. A categorical variable of district, with Mayuge as the reference, was included to distinguish between the Eastern and Western regions of Uganda. The year of study recruitment was added as a categorical variable to capture any unobserved time-dependent effects, where 2022 was the reference.

### Statistical models

All analyses were conducted using R (version 4.2.1) [40]. To model the cured outcome, we only considered individuals who were positive at baseline (1406 individuals; 637 adults and 769 children). We ran logistic regressions for the primary outcomes of being cured without any covariate selection, since covariates were chosen based on the literature. Multicollinearity was checked using the variance inflation factor (VIF) with a cut-off of *>* 5 indicating collinearity [41]. We also ran separate models for adults and children to examine potential differences between age groups. We computed join count statistics [42] to assess the presence of spatial autocorrelation in the cured binary outcome [43]. To establish whether any spatial autocorrelation remained in the residuals of the model, we computed Moran’s *I* [44]. For the overall model, we also performed variable selection using stepwise regression (backward elimination) based on the Bayesian information criterion (BIC) and Akaike information criterion (AIC) to assess model parsimony.

## Results

### S. mansoni cure rates

Among the 3870 clinical participants, 137 did not receive treatment (and 29 participants had missing treatment information). Untreated individuals and participants with missing clinical data were removed to obtain a study population of 3395 treated participants. Fig. 1 shows the study flowchart resulting in 1406 positive participants at baseline diagnosed via KK microscopy and treated. An equivalent figure is included in the supplement for POC-CCA testing (Fig. S1), where 2881 participants had required clinical data. At baseline, the number of participants who were infected was 1990 by POC-CCA trace positive and 1514 by POC-CCA trace negative.

**Fig. 1:**
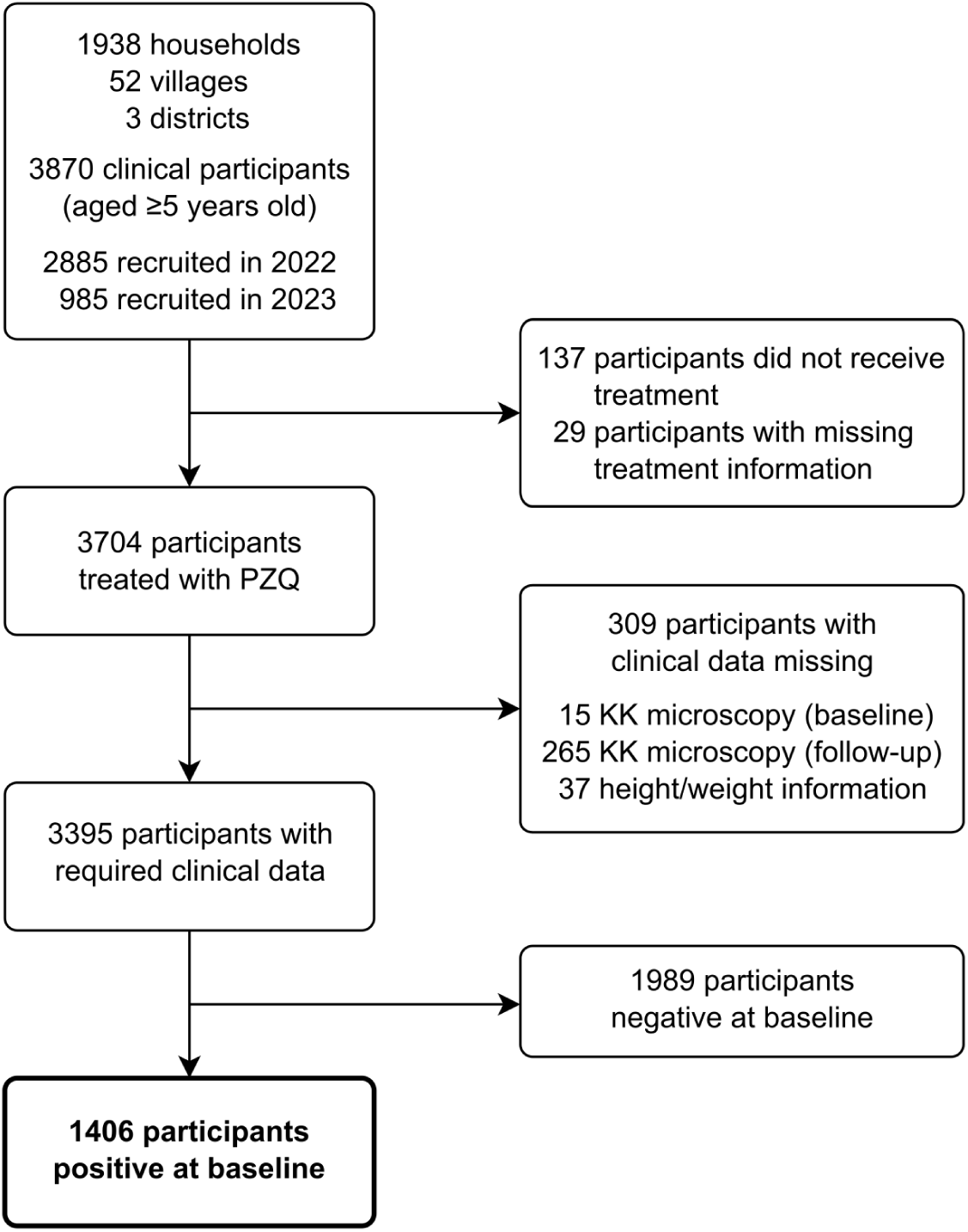
Participant flowchart.

Reasons for no PZQ administration included participants having a history of seizures (6.6%, 9/137), being pregnant (13.9%, 19/137), showing symptoms of severe illness (6.6%, 9/137), or refusing medication (73.0%, 100/137). Over 41% (1406/3395) of participants were infected with *S. mansoni* at baseline, and 14.3% (486/3395) of participants were infected at the drug efficacy follow-up. For positive infections, there was a median of 84 EPG (interquartile range (IQR): [24, 276]) at baseline and 60 EPG (IQR: [12, 192]) at follow-up. The overall cure rate was 76.3% (1073/1406) for ERR = 100%, and 81.9% (1151/1406) for ERR *≥* 90%. Table 1 shows a breakdown of these values by year and district. POC-CCA cure rates were lower than KK cure rates (for both ERR thresholds), with cure at 50.5% (766/1514) for POC-CCA trace negative and 31.7% (631/1990) for POC-CCA trace positive.

**Table 1:** Cure rate summary.

|  | 2022 |  |  |  | 2023 |  |  |
| --- | --- | --- | --- | --- | --- | --- | --- |
|  | Mayuge<br>(N=891) | Buliisa<br>(N=864) | Pakwach<br>(N=877) | Overall<br>(N=2632) | Buliisa<br>(N=209) | Pakwach<br>(N=554) | Overall<br>(N=763) |
| **Baseline infection status** |  |  |  |  |  |  |  |
| negative | 568 (63.7%) | 472 (54.6%) | 434 (49.5%) | 1474 (56.0%) | 156 (74.6%) | 359 (64.8%) | 515 (67.5%) |
| positive | 323 (36.3%) | 392 (45.4%) | 443 (50.5%) | 1158 (44.0%) | 53 (25.4%) | 195 (35.2%) | 248 (32.5%) |
| **Baseline EPG (for positive)** |  |  |  |  |  |  |  |
| Median | 60.0 | 120 | 120 | 96.0 | 36.0 | 60.0 | 60.0 |
| [IQR] | [24.0, 204] | [46.5, 324] | [36.0, 336] | [36.0, 300] | [12.0, 84.0] | [24.0, 204] | [12.0, 192] |
| **Follow-up infection status** |  |  |  |  |  |  |  |
| negative | 831 (93.3%) | 684 (79.2%) | 712 (81.2%) | 2227 (84.6%) | 193 (92.3%) | 489 (88.3%) | 682 (89.4%) |
| positive | 60 (6.7%) | 180 (20.8%) | 165 (18.8%) | 405 (15.4%) | 16 (7.7%) | 65 (11.7%) | 81 (10.6%) |
| **Follow-up EPG (for positive)** |  |  |  |  |  |  |  |
| Median | 42.0 | 48.0 | 54.0 | 48.0 | 48.0 | 24.0 | 36.0 |
| [IQR] | [12.0, 123] | [12.0, 216] | [12.0, 156] | [12.0, 156] | [12.0, 192] | [12.0, 120] | [12.0, 144] |
| **Cure rate\*** | 85.4% | 68.9% | 72.9% | 75.0% | 84.9% | 81.5% | 82.3% |

\* Pairwise comparison of cure rates (100% ERR) across districts using Bonferroni-adjusted proportion tests; significant differences observed between Mayuge and Buliisa ( $P < 0.001$ ) and between Mayuge and Pakwach ( $P < 0.001$ ), but not between Pakwach and Buliisa ( $P = 0.280$ ).

Fig. 2 shows the transitions between no infection and low, moderate, and heavy infection intensities at baseline and follow-up based on KK microscopy. No difference in their infection intensity was observed in 58.3% (1978/3395) of individuals (including those who were negative at both baseline and follow-up). However, 4.1% (140/3395) moved from a heavier to a lower intensity (heavy to moderate or low and moderate to low), indicating some infection clearance. Participants with low baseline infection intensity had a cure rate of 83.0% (628/757). An equivalent plot for POC-CCA testing is shown in Fig. S2, where attempts were made to assess intensity based on test to control line comparisons (see Supplementary methods). There was a lower percentage of participants having the same intensity at baseline and follow-up (39.8%, 1147/2881), while 7.5% (217/2881) of participants moved from a heavier to a lower intensity, for POC-CCA trace negative and 20.6% (594/2881) for POC-CCA trace positive. Fig. S3 shows the agreement between POC-CCA and KK tests at baseline and follow-up as confusion matrices. When trace was considered as negative, comparing the distributions of baseline infection intensities between the two tests using a Chi-squared test showed a significant difference (*χ*^2^ = 163, df = 3*, P <* 0.001). Similarly, when trace was considered as positive (low intensity, i.e. here when the treatment line was fainter than the control line), the difference between the distributions was still significant (*χ*^2^ = 653, df = 3*, P <* 0.001). Considering the 3017 individuals who had KK microscopy and POC-CCA results at baseline, agreement was computed using weighted Cohen’s kappa with a value of 0.37 (95% confidence interval (CI): 0.30 to 0.44) when trace was taken as negative and 0.35 (95% CI: 0.29 to 0.40) when trace was taken as positive (low intensity), showing fair agreement. However, POC-CCA classified 4.9% (149/3017) of participants as negative and 5.2% (157/3017) as trace, despite being positive by KK microscopy.

**Fig. 2:**
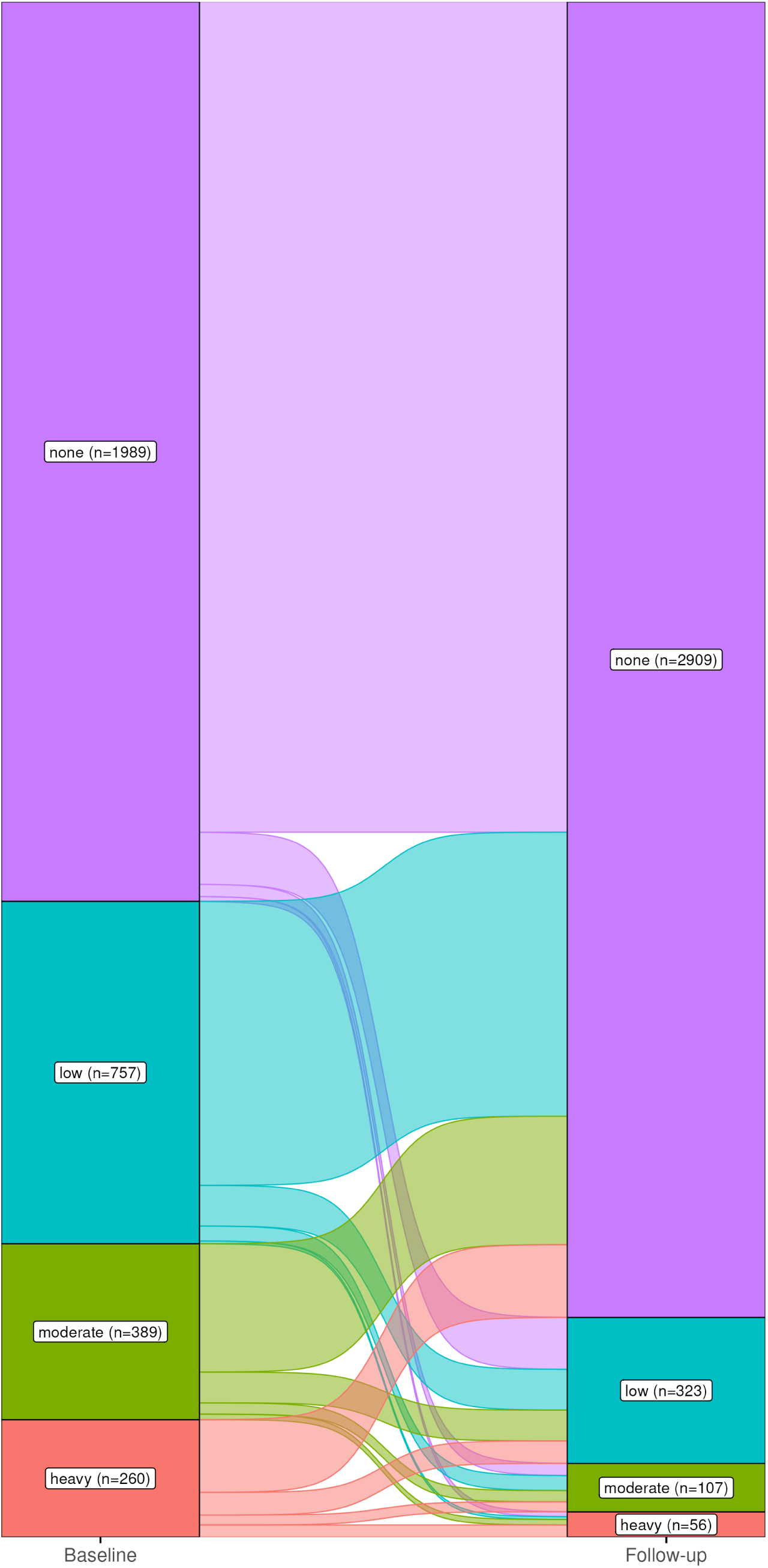
Kato–Katz baseline to follow-up. 4.5% (153/3395) of individuals were negative at baseline and positive at follow-up. 1.5% (51/3395) of individuals moved from a lower to a higher intensity.

### Determinants of cure from praziquantel

Table 2 shows the summary of all covariates considering the study population of 3395 treated participants (with required clinical data) and the 1406 positive (by KK) participants.

**Table 2:** Summary table of participant characteristics.

| Covariate | All participants<br>(*n* = 3395) | Positive participants<br>(*n* = 1406) | *p*-value |
| --- | --- | --- | --- |
| **Sociodemographic** |  |  |  |
| Age (median [IQR]) | 17.0 [9.0, 38.0] | 15.0 [10.0, 33.0] | 0.16 |
| Gender (%) |  |  | <0.01 |
| Male | 1580 (46.5) | 722 (51.4) |  |
| Female | 1815 (53.5) | 684 (48.6) |  |
| Educational attainment (median [IQR]) | 3.0 [1.0, 5.0] | 3.0 [1.0, 5.0] | 0.24 |
| Occupation (%) |  |  | 0.03 |
| None/other | 3036 (89.4) | 1225 (87.1) |  |
| Fishing | 248 (7.3) | 135 (9.6) |  |
| Fishmongering | 111 (3.3) | 46 (3.3) |  |
| Household years in village (median [IQR]) | 16.0 [8.0, 30.0] | 15.0 [7.0, 27.0] | 0.26 |
| **Biomedical** |  |  |  |
| Received PZQ in the past year (%) |  |  | 0.65 |
| No | 1763 (51.9) | 741 (52.7) |  |
| Yes | 1632 (48.1) | 665 (47.3) |  |
| Height (median [IQR]) | 153.0 [130.0, 164.0] | 154.0 [134.0, 164.0] | 0.12 |
| Weight (median [IQR]) | 46.0 [26.0, 58.1] | 45.3 [27.6, 57.1] | 0.54 |
| **WASH** |  |  |  |
| Improved sanitation (%) | 1926 (56.7) | 799 (56.8) | 0.98 |
| **Water contact** |  |  |  |
| Domestic activities (%) | 1216 (35.8) | 485 (34.5) | 0.40 |
| Occupational activities (%) | 592 (17.4) | 289 (20.6) | 0.01 |
| Recreational activities (%) | 124 (3.7) | 69 (4.9) | 0.05 |
| **Spatial** |  |  |  |
| Distance to nearest health centre (in km)<br>(median [IQR]) | 2.4 [1.5, 4.1] | 2.5 [1.7, 4.0] | 0.15 |
| Distance to closest site with snails (in m)<br>(median [IQR]) | 335.7 [169.3, 722.2] | 287.0 [140.5, 631.0] | <0.01 |
| District (%) |  |  | 0.04 |
| Mayuge | 891 (26.2) | 323 (23.0) |  |
| Buliisa | 1073 (31.6) | 445 (31.7) |  |
| Pakwach | 1431 (42.2) | 638 (45.4) |  |
| **Study design** |  |  |  |
| Year of recruitment (%) |  |  | <0.01 |
| 2022 | 2632 (77.5) | 1158 (82.4) |  |
| 2023 | 763 (22.5) | 248 (17.6) |  |

Notes: For binary and other categorical covariates, counts and percentages are reported [*n* (%)], and group differences were evaluated using a chi-squared test. For all non-normally distributed covariates, results are summarised as median and IQR (median [IQR]), and group differences were evaluated using the Wilcoxon rank-sum test.

Fig. 3 shows the model of cure rate for ERR = 100%. Each one-year increase in age was associated with 1.03 times higher odds of being cured (95% CI: 1.02 to 1.04). Equivalently, each 10-year increase in age was associated with 1.34 times higher odds of being cured (95% CI: 1.17 to 1.53). Participants with moderate or heavy baseline infections had lower odds of being cured compared with those with light infections (OR range 0.39 to 0.61). Past PZQ treatment through MDA was an insignificant predictor of cure. Furthermore, each one-centimetre increase in height was associated with 0.01 lower odds of being cured (OR 0.99, 95% CI: 0.98 to 1.00), and equivalently each 10 cm increase was associated with 0.12 lower odds of being cured (OR 0.88, 95% CI: 0.79 to 0.98). Participants in the Western districts of Buliisa and Pakwach had lower odds (OR range 0.48 to 0.53) of being cured compared with those in the Eastern district of Mayuge.

**Fig. 3:**
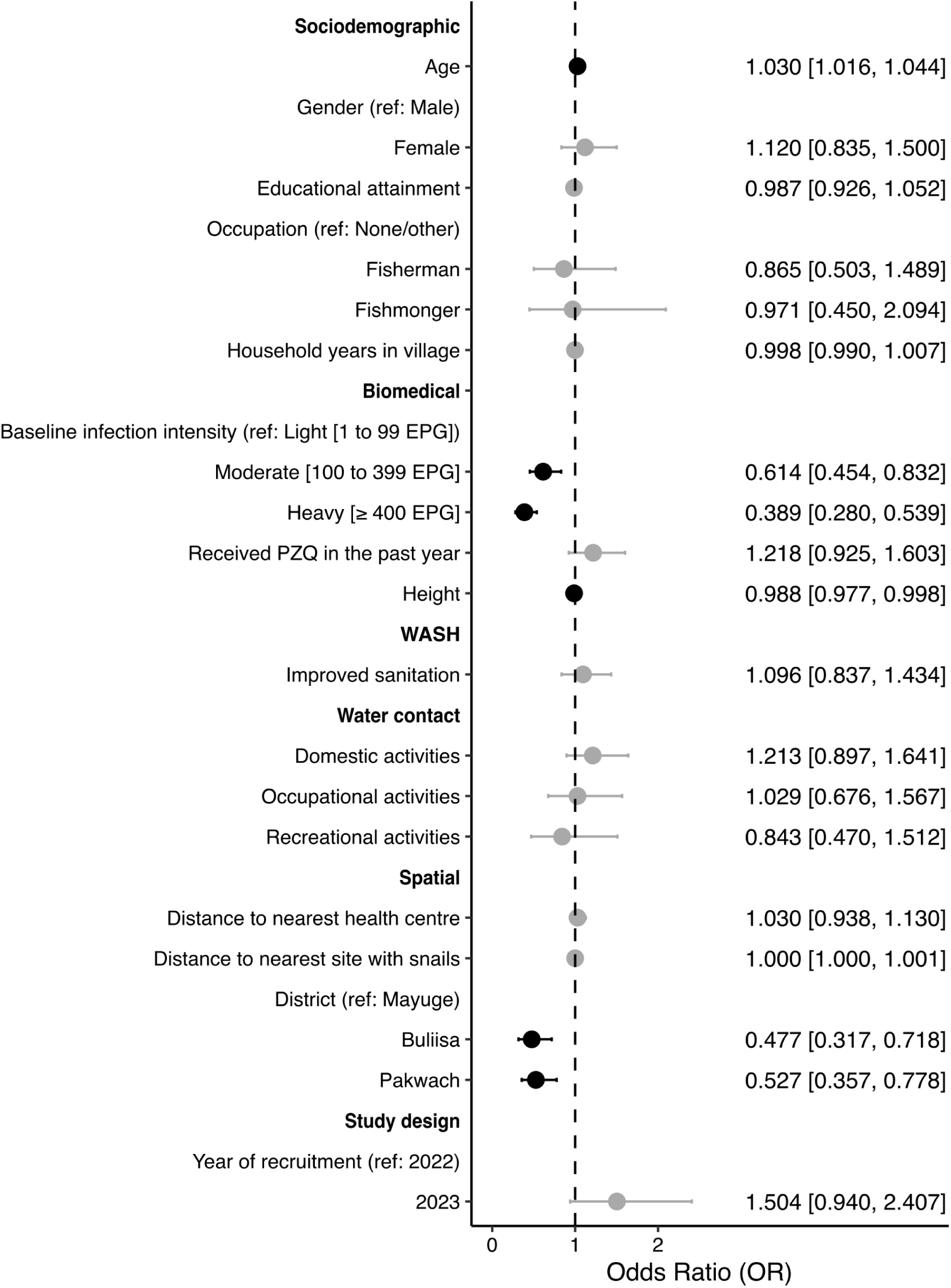
Determinants of being cured after treatment. The outcome was 100% egg reduction rate as determined through Kato–Katz microscopy. ORs (exponentiated coefficient estimates represented by the dots) of significant (*P <* 0.05) coefficients are shown in black, and of non-significant coefficients in grey. 95% CIs are indicated in lines. OR and CI values are also reported. Height and weight were collinear; therefore, weight was removed.

Sub-group analyses of adults and children are presented in Fig. 4. In the adults-only model, the same coefficients as the full model were significant. Notably, each one-year increase in age was associated with 1.05 times higher odds of being cured (95% CI: 1.03 to 1.07), compared to 1.03 in the full model, and each one-centimetre increase in height was associated with 0.03 lower odds of being cured (OR 0.97, 95% CI: 0.94 to 1.00). These two covariates were insignificant in the children-only model. For every 100-metre increase in distance to the nearest water site with snail presence, there was 1.05 (95% CI: 1.00 to 1.12) times higher odds of being cured in the children-only model (*p*-value = 0.04). Unlike in the adults-only model, there was no observed difference between the districts of Buliisa and Mayuge for children only, but the significant difference between Pakwach and Mayuge, which was present in the full model, remained.

**Fig. 4:**
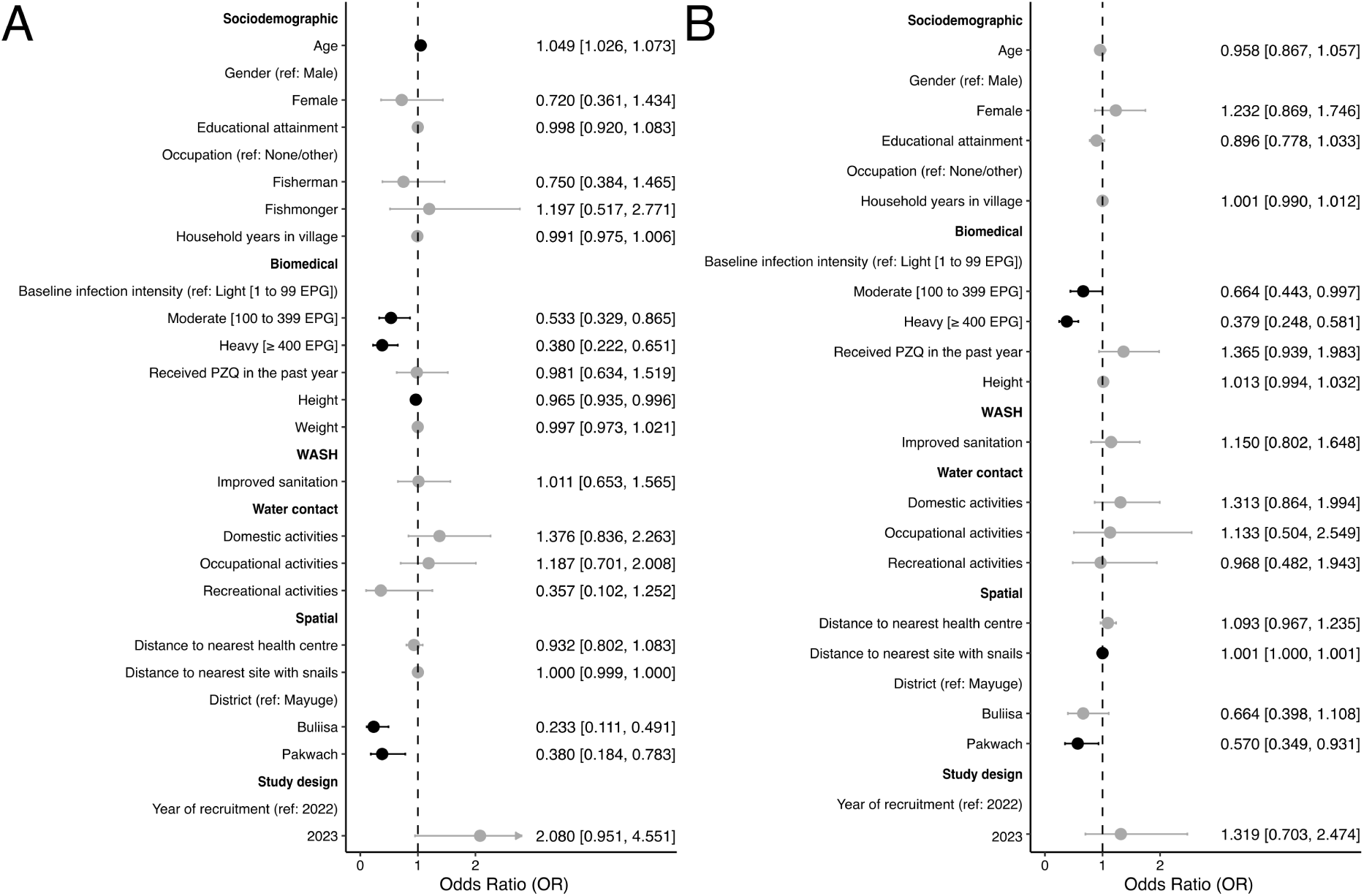
Adults- and children-only models. Determinants of being cured after treatment based on 100% egg reduction rate as determined through Kato–Katz microscopy. Panel **A** shows the adults-only model, and panel **B** shows the children-only model. ORs (exponentiated coefficient estimates represented by the dots) of significant (*P <* 0.05) coefficients are shown in black, and of non-significant coefficients in grey. 95% CIs are indicated in lines. OR and CI values are also reported. Weight was not excluded in the adults-only model as no collinearity was observed with height, but it was excluded in the children-only model due to collinearity. Occupation covariate excluded in children-only model due to insufficient variation (less than 1% of children reported an occupation other than the reference category).

A total of 17.1% (241/1406) of participants who were positive at baseline lay on the boundary of their respective dosage category. We visualised the height distributions of children and adults based on the dosage administration categories (Fig. S4). From BMI calculations, we observed that 14.6% (93/637) of adults were classified as underweight, 69.4% (442/637) as normal, 11.9% (76/637) as overweight (but not obese), and 4.1% (26/637) as obese. Furthermore, when calculating the number of tablets that an individual should have received as per the 40 mg*/*kg recommended dosage, and compared to the number of tablets they received based on the dose pole, we observed 39.6% (557/1406) of all participants having been underdosed, with this number being 57.1% (364/637) for adults only.

### Sensitivity and robustness analyses

Stepwise variable selection using AIC and BIC produced more parsimonious models (resulting in a total of six and five covariates, respectively; Figs. S5 and S6), however the magnitudes of the significant coefficients remained largely similar to those in the full model. The model using a *≥* 90% ERR to define cure (Fig. S7) had the same significant coefficients as the 100% ERR model, except that baseline infection intensity showed no significant difference between moderate and heavy intensities compared with low intensity. Both POC-CCA models (Figs. S8 and S9) had age, height, and baseline infection intensity as significant coefficients, similar to the full model. The district effect was lost, with only Pakwach being significantly different from Mayuge in the model where trace was considered negative. In the POC-CCA trace positive model, fishermen had higher odds (OR 1.71, 95% CI: 1.08 to 2.71) of being cured compared to the reference category of none/other occupation, and household years in village was also significant with higher odds (OR 1.00, 95% CI: 1.00 to 1.02) of cure with every one-year increase. In the POC-CCA trace negative model, distance to the nearest health centre from the household of a participant was associated with 1.09 (95% CI: 1.01 to 1.17) higher odds of being cured for every one-kilometre increase. Year of recruitment was significant in both models, with 2023 having lower odds of cure (OR range 0.30 to 0.45) compared to 2022.

### Spatial autocorrelation

Both cured and non-cured participants were spatially clustered by district rather than randomly distributed. The join count test detected significant positive spatial autocorrelation within districts for treatment outcomes in both cured and non-cured participants. For non-cured individuals, the number of same-category joins (observed = 41.22) was significantly higher than that expected (expected = 39.34) under spatial randomness (*Z* = 10.37, *P <* 0.001). Cured individuals showed an equivalent pattern (observed = 411.22, expected = 409.34; *Z* = 10.37, *P <* 0.001). No significant spatial autocorrelation was detected in the model residuals when Moran’s *I* was calculated (*I* = *−*0.001, *p*-value = 0.6). However, when the district effect was removed, the residuals showed significant positive spatial autocorrelation (*I* = 0.008, *p*-value *<* 0.001), suggesting that the district effect accounted for most of the spatial dependence.

## Discussion

Variation in PZQ efficacy is common and, if formally monitored in endemic countries, may help inform strategies to optimise MDA campaigns. In this study, we tested and treated a total of 3704 adult and child participants living in fishing communities in rural Uganda during two field surveys conducted in 2022 and 2023, as part of the SchistoTrack cohort. Among a diverse set of sociodemographic, biomedical, WASH, water contact, spatial, and other study design factors, individual-level and spatial factors primarily determined cure.

The overall cure rate of 76.3% observed in our study, based on KK microscopy, is within the very large variability of PZQ efficacy previously shown in East Africa, ranging from 61.8 to 99.1% [4]. Cure rates varied by district, from 70.8% in Buliisa, to 75.5% in Pakwach, and 85.4% in Mayuge. Spatial autocorrelation in the cured outcome prior to modelling confirmed the presence of spatial heterogeneity in PZQ efficacy. Including the district covariate as a fixed effect adequately controlled for this dependence, but nonetheless indicated significant differences in treatment outcomes between the Western and Eastern districts of Uganda. Despite the inclusion of detailed sociodemographic, biomedical, and WASH covariates capturing individual- and household-level factors, there remained substantial unexplained geographical variation in the odds of being cured. Previous studies have reported higher prevalence and morbidity related to *S. mansoni* infections in the areas near Lake Albert (Western Uganda) [11, 12], suggesting that factors beyond individual characteristics, such as local environmental conditions or parasite population structure, may influence treatment outcomes. These findings strengthen the need to incorporate spatial structure into future statistical or mathematical models of transmission, drug response, and control, as assuming uniform efficacy across regions risks overlooking important geographic trends and could lead to misleading inferences and suboptimal control strategies.

Age was a significant predictor of being cured, with higher odds among older participants. When analysed separately, age was not significant in the children-only model, consistent with studies in the literature that focused on preschool and/or school-aged children [3, 9, 10, 18–20]. Sousa-Figueiredo et al. found differences in cure rates between age groups of 1–3 and 4–7 years old [16], but our study did not include children younger than 5 years old. On the other hand, age remained a significant predictor for adults only, contrasting other studies that reported no association between cure rates and age [14, 45]. These findings suggest that age may influence treatment response differently across demographic groups, potentially reflecting cumulative exposure, acquired immunity, or differences in drug metabolism. Historical adherence to treatment coupled with different sanitary practices between adults and children may also influence the development of acquired immunity over time [46]. practices The significance of distance to the nearest water site with snail presence among children but not adults further supports this hypothesis, indicating that current exposure may play a stronger role in determining cure rates for children than for adults.

Height, used to determine the administration of PZQ during MDA by creating height categories in the WHO dose pole, also was a significant predictor in adults, with taller individuals showing lower odds of being cured. Interestingly, weight, which was not collinear with height for adults, was insignificant. This pattern could perhaps indicate underdosing of taller individuals or among individuals close to the upper boundaries of the height-based dose categories. It also may reflect that the height-to-weight approximation used by the WHO for the dose pole needs to be updated. A recent systematic review by Berry et al. [35] found that using the dose pole for treatment administration results in nearly 20% of the adult population being underdosed, likely due to increased obesity observed since the dose pole was developed. Surprisingly, a notable proportion of individuals in our study were classified as overweight based on BMI, consistent with reports of rising overweight and obesity rates contributing to the emergence of non-communicable diseases in rural areas of sub-Saharan Africa [47, 48]. The systematic review by Berry et al. [35] did not find this inaccurate dosage in child populations, and this trend was not present in our children-only model either, though without clear explanation, Crellen et al. [17] suggested that lower weight in children was associated with lower ERRs. Whether these relationships were due to suboptimal dosing or other factors, such as pharmacokinetic variation or behavioural exposure differences, remains unclear, and future work is necessary, especially focused on correct dosing in adults.

There is no agreement as to whether participation in multiple MDA rounds has a positive or negative impact on PZQ efficacy, especially in the context where PZQ resistance is unclear. Crellen et al. [17] reported lower ERRs with higher MDA exposure, whereas Walker et al. [10] discuss hypotheses that repeated treatments might lead to stronger acquired immunity and improve efficacy in subsequent treatments. Immunological studies support this latter mechanism, with evidence showing that repeated exposure to *S. mansoni* antigens released during PZQ-induced worm death can boost protective antibody responses [49–51], though the extent to which this translates into improved PZQ efficacy remains unresolved. In our models, receiving PZQ within the past year through MDA showed a positive but insignificant relationship with being cured. Age, used alongside household years in village as a proxy for past rounds of MDA, was significantly associated with cure. District differences also indicated more favourable cure rates in the Eastern district of Mayuge, which has received more rounds of MDA (15 since 2003), compared to the Western districts of Buliisa and Pakwach (13) [11]. These patterns provide some support for the hypothesis that acquired immunity contributes to improved PZQ efficacy. Future studies integrating genomic surveillance of *S. mansoni* populations and pharmacokinetic assessments could help differentiate between true resistance, reduced sensitivity, and dosing or diagnostic artefacts, thus longitudinal designs and routine monitoring of PZQ efficacy following MDA campaigns are essential.

Discrepancies between KK and POC-CCA results highlight diagnostic limitations in evaluating PZQ efficacy. The KK method, while specific, may underestimate residual infection due to low sensitivity at light intensities, whereas POC-CCA may overestimate infection persistence due to antigen clearance lag or cross-reactivity [52–54]. These methodological differences likely explain the lower cure rates when using POC-CCA, also observed by Kildemoes et al. [55]. Measurement error and diagnostic uncertainty must be considered when interpreting apparent spatial or demographic heterogeneity in treatment response. In our POC-CCA models, the recruitment year covariate was taken as a proxy for the different batches used in the two surveys, and the results showed a significant difference between them, agreeing with previous studies that POC-CCA tests show wide batch-to-batch variability [56].

Our study included a comprehensive set of variables with detailed individual and household-level data from a large sample of individuals spanning a wide age range across three districts with differing treatment responses. However, several limitations should be noted. We could not account for potential unmeasured confounders such as nutritional status, and we did not investigate mechanisms of parasite clearance (e.g. pharmacokinetics or metabolism). Diagnostic variability between KK and POC-CCA introduces some uncertainty in cure rate estimates, and the cross-sectional design limited our ability to explore temporal variation in treatment response.

## Conclusion

Current WHO guidelines for PZQ administration have not been substantially revised despite improved evidence on PZQ dose–response relationships and variable cure rates from PZQ across settings. Fixed-dose strategies and uniform efficacy assumptions may no longer reflect field realities. Our study provides evidence that both individual and spatial-level factors influence PZQ efficacy against *S. mansoni* in endemic communities in Uganda. Future control strategies would benefit from integrating spatially explicit analyses, more precise dosing assessments, and consistent post-MDA monitoring to detect shifts in efficacy. Regular evaluation of diagnostic tools and dose pole performance should accompany these efforts. Updating WHO treatment guidelines to reflect contemporary data on dosing accuracy, population diversity, and local transmission dynamics may help maintain PZQ effectiveness and ensure the long-term sustainability of schistosomiasis control programmes.

## Declarations

### Ethics approval and consent to participate

Data collection and use were reviewed and approved by the Oxford Tropical Research Ethics Committee (OxTREC 509-21), the Vector Control Division Research Ethics Committee of the Uganda Ministry of Health (VCDREC146), and the Uganda National Council for Science and Technology (UNCST HS1664ES). Written informed consent was obtained from all adults, who also consented on behalf of those under 18 years of age. All children provided informed verbal assent.

### Consent for publication

Not applicable.

### Data availability

The datasets generated and analysed during the study are not publicly available due to the identifiable and sensitive nature of the participant characteristics and the ongoing status of the SchistoTrack cohort. Relevant metadata are included in the article or the additional files.

### Author contributions

Conceptualisation: MAI, FR, and GFC. Data curation: MAI, FR, SW, AE, EA, JN, AN, MS, BN, NBK, and GFC. Formal analysis: MAI, FR, and SW. Funding acquisition: GFC. Investigation: MAI, FR, SW, and GFC. Methodology: MAI, FR, SW, and GFC. Project administration: NBK and GFC. Resources: NBK and GFC. Software: GFC. Supervision: GFC. Validation: MAI, FR, and SW. Visualisation: MAI. Writing—original draft: MAI and GFC. Writing—review & editing: MAI, FR, SW, AE, EA, JN, AN, MS, BN, NBK, and GFC.

## Acknowledgements

We thank the members of the SchistoTrack group for their valuable feedback and insights. We also thank all field teams, specifically surveyors, technicians, nurses, and auxiliary workers. A special acknowledgement is due to the study participants, village health team members, local government nurses, and district leadership.

