## Supplementary methods for "Individual and spatial heterogeneity of praziquantel efficacy against *Schistosoma mansoni* within the context of repeated mass drug administration"

### Supplementary information

#### Supplementary methods

##### POC-CCA testing

Point-of-care circulating cathodic antigen (POC-CCA) tests (supplier: ICT International; product number: SCH25; lot number: 210811080 in 2022 and 221117133 in 2023) were also used to determine infection status. Technicians prepared the test cassettes by transferring two drops of urine to the circular well of the cassette from a well-mixed urine sample provided by the participants. Results were read approximately 20 minutes after preparation. A positive test was recorded if both the control and test lines appeared. A negative test was recorded if only the control line appeared. A test was considered invalid if only the treatment line appeared or if the urine did not flow up to the test band, in which case the test was repeated up to three times before recording an invalid result. The results were classified based on a test–control line comparison, where pos3 indicated a much darker test line than the control line, pos2 indicated a similar test line to the control line, pos1 indicated a fainter test line than the control line, and trace indicated a barely visible test line.

### Supplementary figures

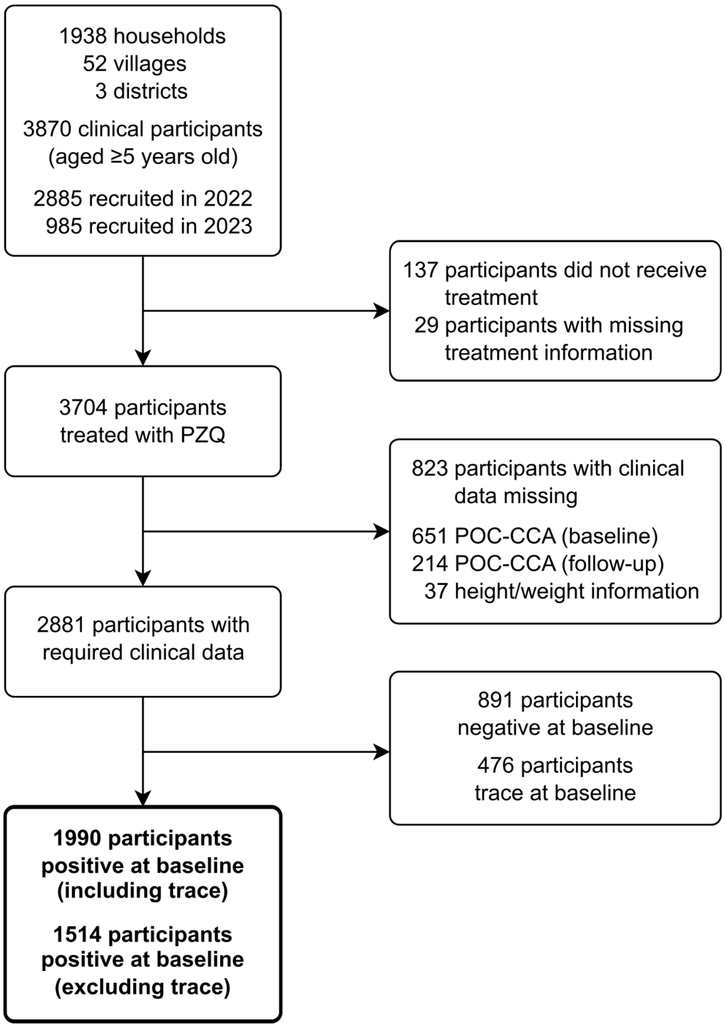


**Fig. S1**: **Participant flowchart (POC-CCA).**

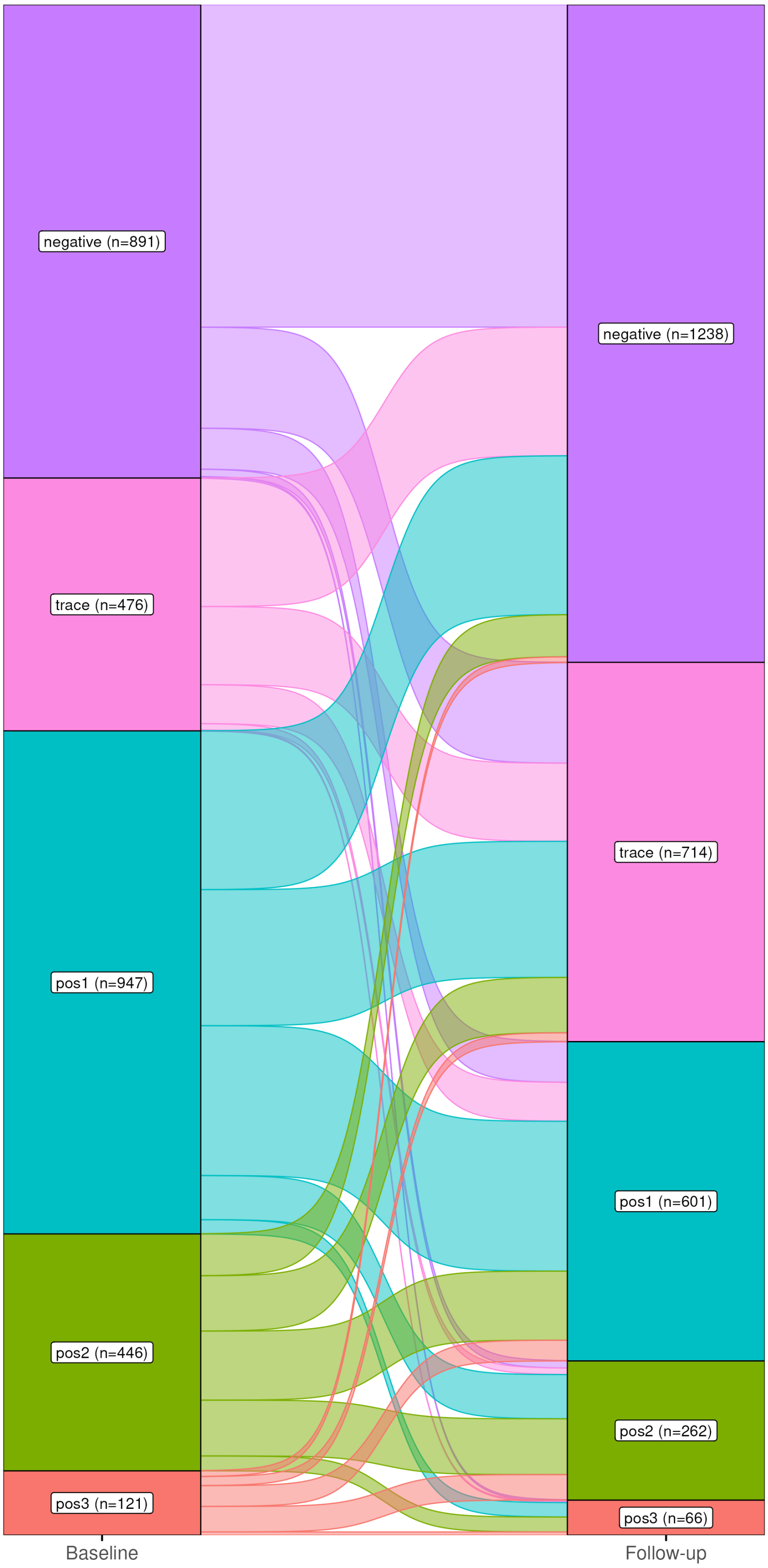


**Fig. S2**: **POC-CCA baseline to follow-up.** 6.3% (181/2881) of individuals were negative at baseline and positive at follow-up, when trace was considered as negative, with 9.9% (284/2881) when trace was considered as positive. 4.8% (138/2881) moved from a lower to a higher intensity when trace was considered negative, and 7.8% (225/2881) when trace was considered positive.

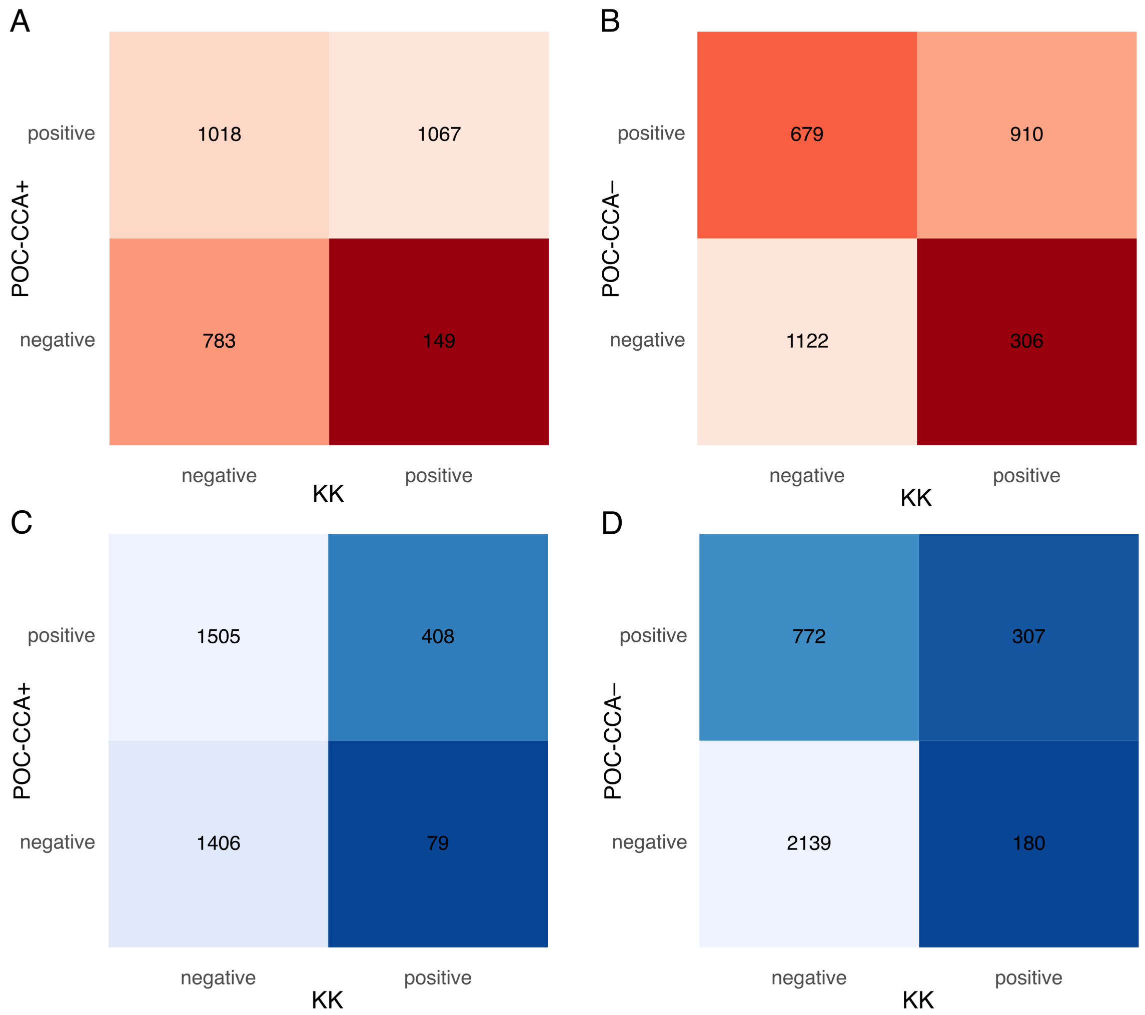


**Fig. S3**: **POC-CCA and Kato–Katz agreement. A, B** Show the agreement between POC-CCA (trace positive and trace negative, respectively) and KK at baseline ($n = 3017$), while **C, D** show the agreement at follow-up ($n = 3398$). Total numbers indicate the number of participants with available information out of the 3704 treated participants.

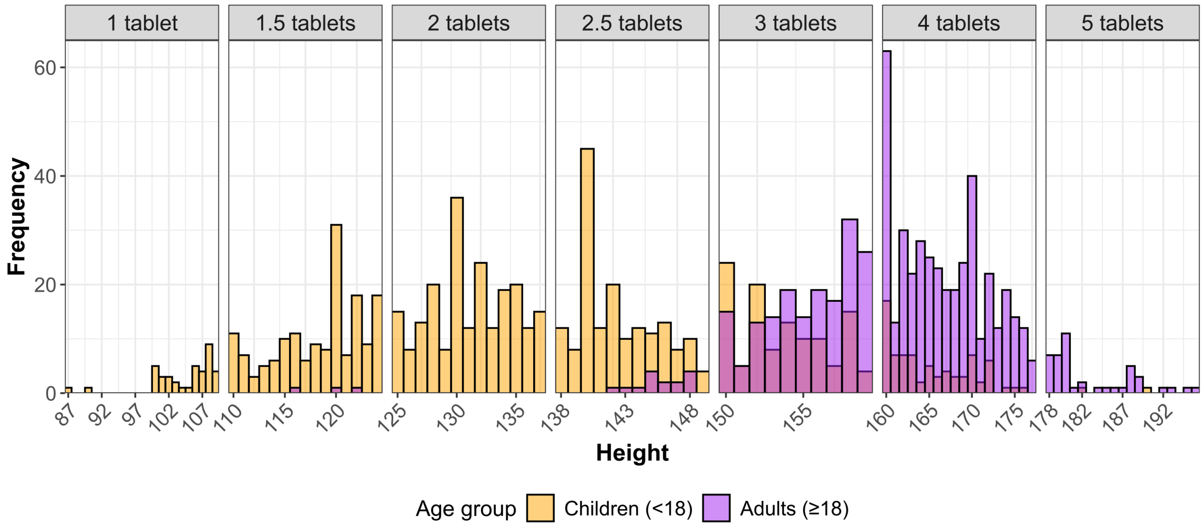


**Fig. S4**: **Height distribution of participants.** The distribution is visualised by age-group, with orange indicating children and purple indicating adults. Bin-widths are set at 1 cm. Each box represents a dosage category based on the WHO's dose pole, as follows: 1 tablet for $\leq$ 109 cm, 1.5 tablets for 110 to 124 cm, 2 tablets for 125 to 137 cm, 2.5 tablets for 138 to 149 cm, 3 tablets for 150 to 159 cm, 4 tablets for 160 to 177 cm, and 5 tablets for $\geq$ 178 cm.

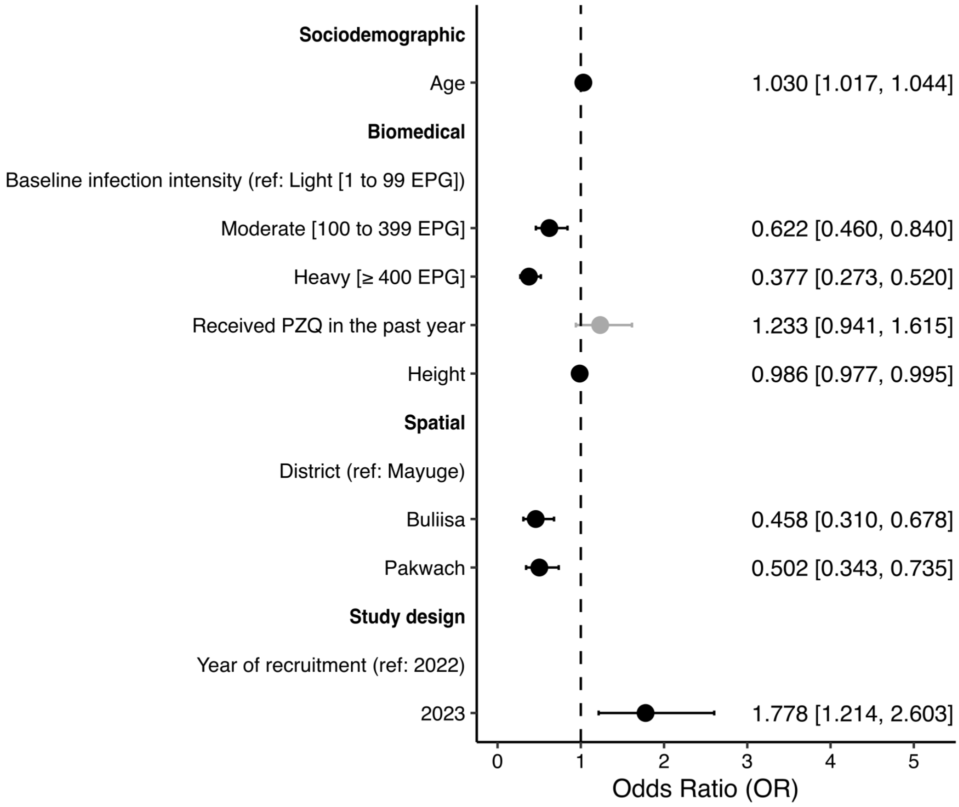


**Fig. S5**: **Determinants of being cured for AIC-based model.** Variable selection by step-wise backwards AIC. ORs (exponentiated coefficient estimates represented by the dots) of significant ($P < 0.05$) coefficients are shown in black, and of non-significant coefficients in grey. 95% CIs are indicated in lines. OR and CI values are also reported.

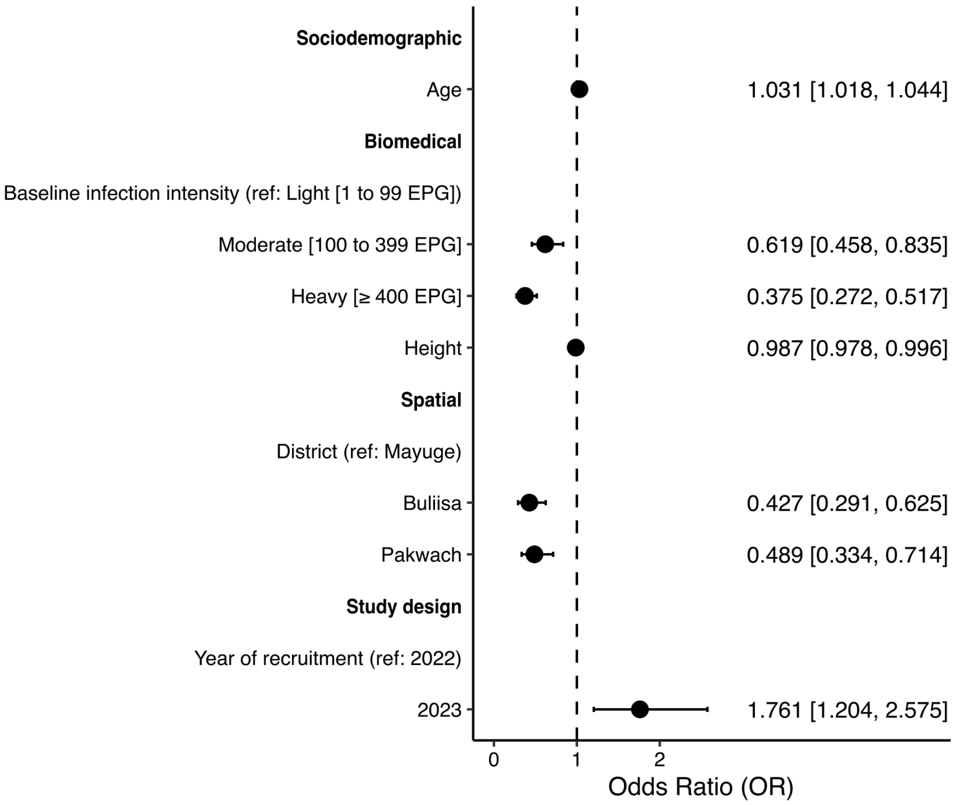


**Fig. S6**: **Determinants of being cured for BIC-based model.** Variable selection by step-wise backwards BIC. ORs (exponentiated coefficient estimates represented by the dots) of significant ($P < 0.05$) coefficients are shown in black, and of non-significant coefficients in grey. 95% CIs are indicated in lines. OR and CI values are also reported.

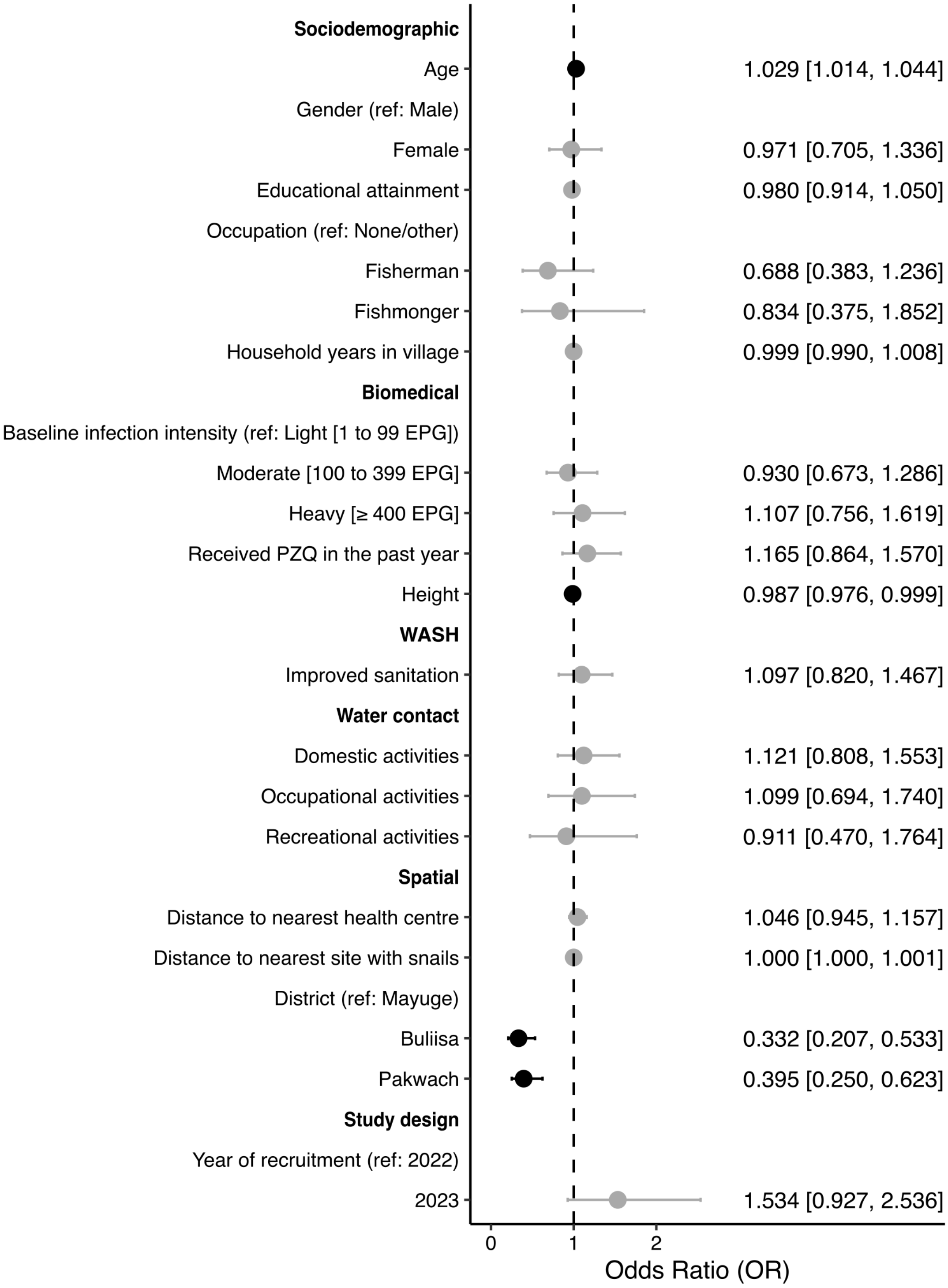


**Fig. S7**: **Determinants of being cured after treatment based on $\geq 90\%$ egg reduction rate.** Determined through Kato–Katz microscopy. ORs (exponentiated coefficient estimates represented by the dots) of significant ($P < 0.05$) coefficients are shown in black, and of non-significant coefficients in grey. 95% CIs are indicated in lines. OR and CI values are also reported.

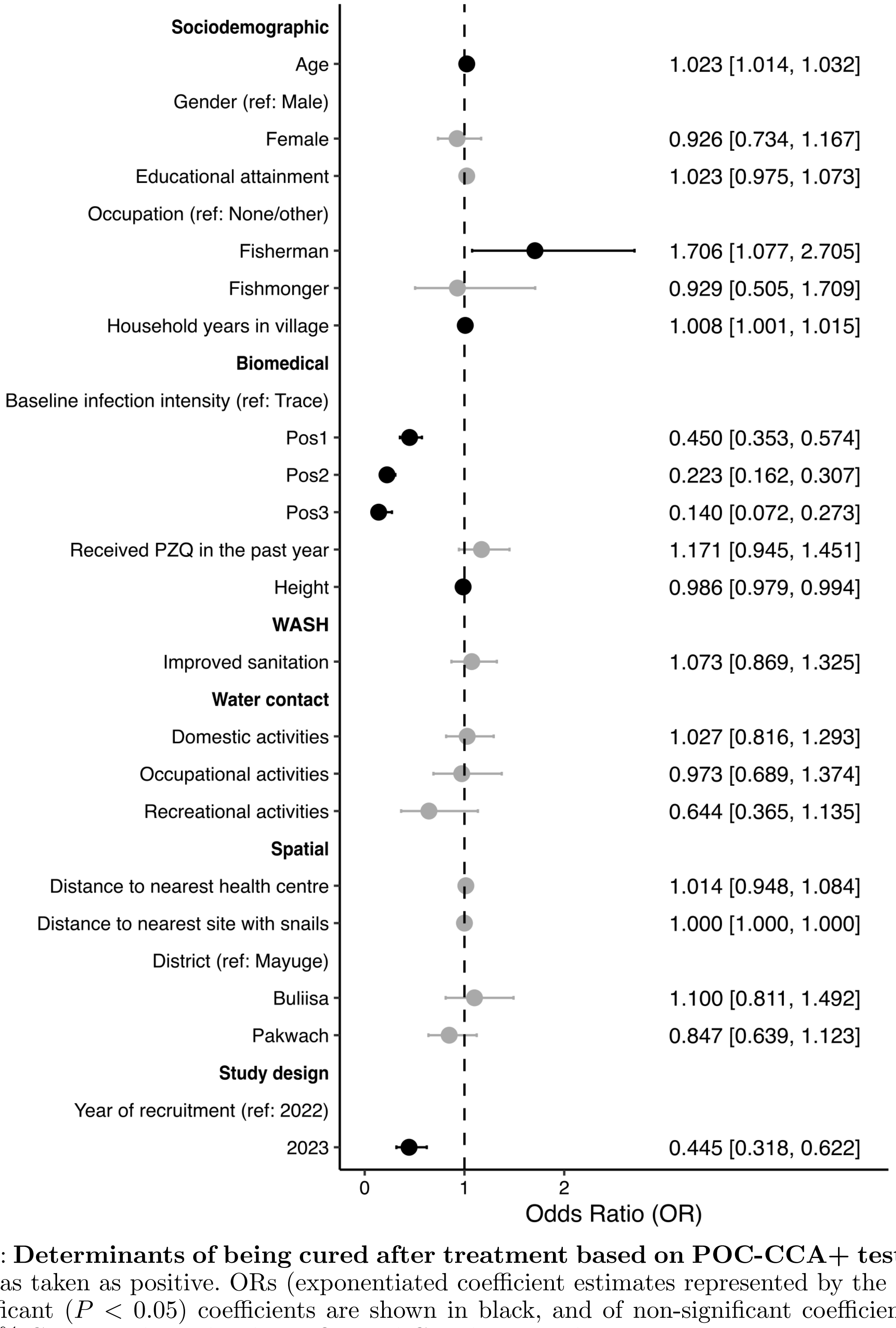


**Fig. S8**: **Determinants of being cured after treatment based on POC-CCA+ testing.** Trace was taken as positive. ORs (exponentiated coefficient estimates represented by the dots) of significant ($P < 0.05$) coefficients are shown in black, and of non-significant coefficients in grey. 95% CIs are indicated in lines. OR and CI values are also reported.

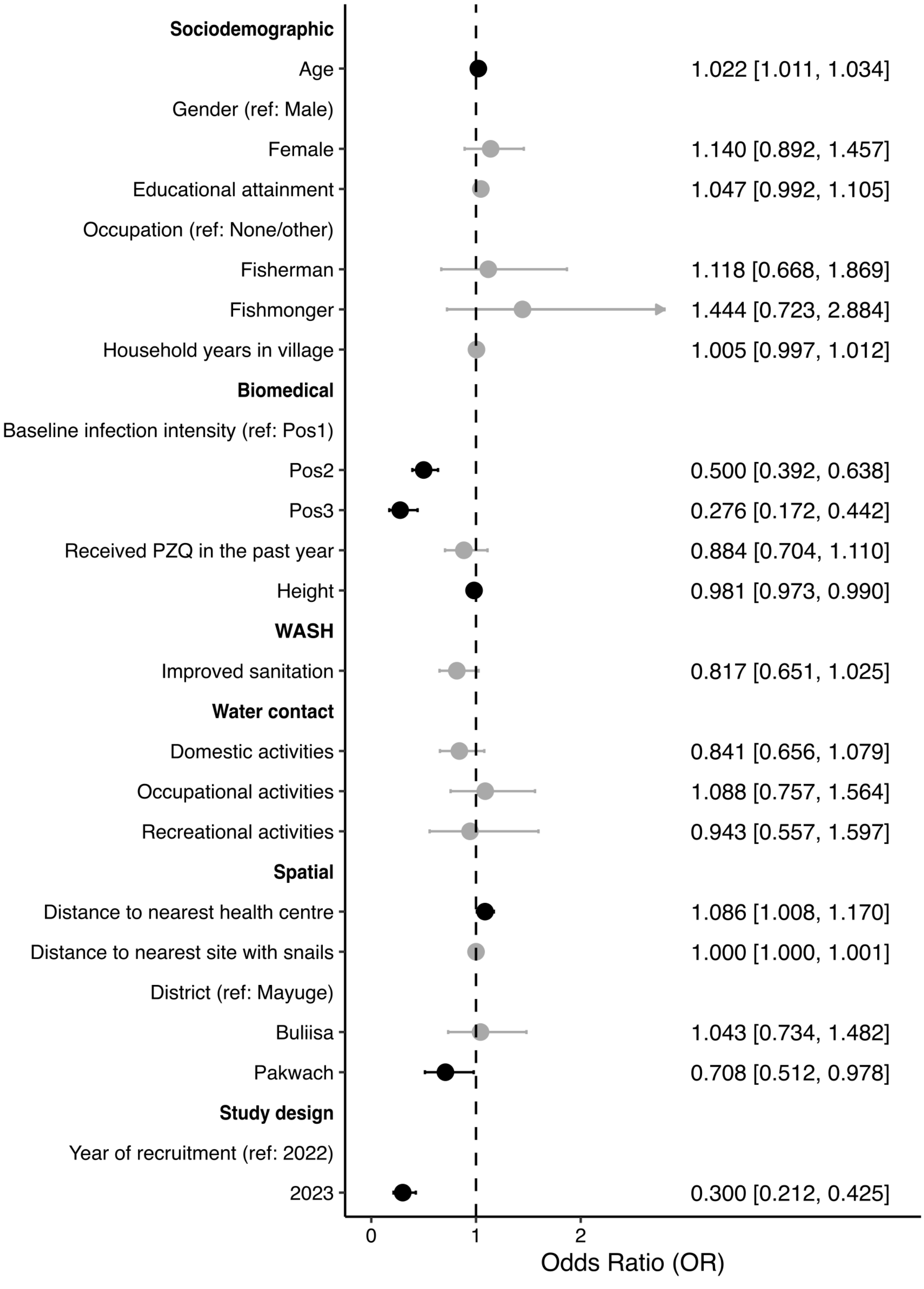


**Fig. S9**: **Determinants of being cured after treatment based on POC-CCA− testing.** Trace was taken as negative. ORs (exponentiated coefficient estimates represented by the dots) of significant ($P < 0.05$) coefficients are shown in black, and of non-significant coefficients in grey. 95% CIs are indicated in lines. OR and CI values are also reported.
